# Aberrant hierarchical prediction errors are associated with transition to psychosis: A computational single-trial analysis of the mismatch negativity

**DOI:** 10.1101/2022.12.20.22283712

**Authors:** Daniel J. Hauke, Colleen E. Charlton, André Schmidt, John Griffiths, Scott W. Woods, Judith M. Ford, Vinod H. Srihari, Volker Roth, Andreea O. Diaconescu, Daniel H. Mathalon

## Abstract

**Background:** Mismatch negativity (MMN) reductions are among the most reliable biomarkers for schizophrenia and have been associated with increased risk for conversion to psychosis in individuals at clinical high risk for psychosis (CHR-P). Here, we adopt a computational approach to develop a mechanistic model of MMN reductions in CHR-P individuals and patients early in the course of schizophrenia (ESZ).

**Methods:** Electroencephalography (EEG) was recorded in 38 CHR-P individuals (15 converters), 19 ESZ patients (≤5 years), and 44 healthy controls (HC) during three different auditory oddball MMN paradigms including 10% duration-, frequency-, or double-deviants, respectively. We modelled sensory learning with the hierarchical Gaussian filter and extracted precision-weighted prediction error trajectories from the model to assess how the expression of hierarchical prediction errors modulated EEG amplitudes over sensor space and time.

**Results:** Both low-level sensory and high-level volatility precision-weighted prediction errors were altered in CHR-P and ESZ groups compared to HC. Furthermore, low-level precision-weighted prediction errors were significantly different in CHR-P that later converted to psychosis compared to non-converters.

**Conclusions:** Our results implicate altered processing of hierarchical prediction errors as a computational mechanism in early psychosis consistent with predictive coding accounts of psychosis. This computational model appears to capture pathophysiological mechanisms relevant to early psychosis and the risk for future psychosis in CHR-P individuals, and may serve as a predictive biomarker and mechanistic target for novel treatment development.

## Introduction

Often without our awareness, our brain continuously learns about the environment that surrounds us. The mismatch negativity (MMN) is a neurophysiological index of such implicit learning commonly measured with electroencephalography (EEG). It refers to a brain response that is elicited automatically when an auditory stimulus violates a statistical regularity in the recent auditory environment,^1^ for example when a series of low tones is unexpectedly interrupted by a high tone (Figure 1A). Formally, the MMN is a transient negative wave deflection in the event-related potential (ERP) elicited by infrequent auditory *deviant* stimuli randomly interspersed among frequent *standard* stimuli that is most easily identified between 100-250ms following stimulus onset in the deviant-standard ERP difference wave (Figure 1B). ^2,3^ Importantly, this neurophysiological assessment is very feasible in a wide range of clinical settings.

**Figure 1.**
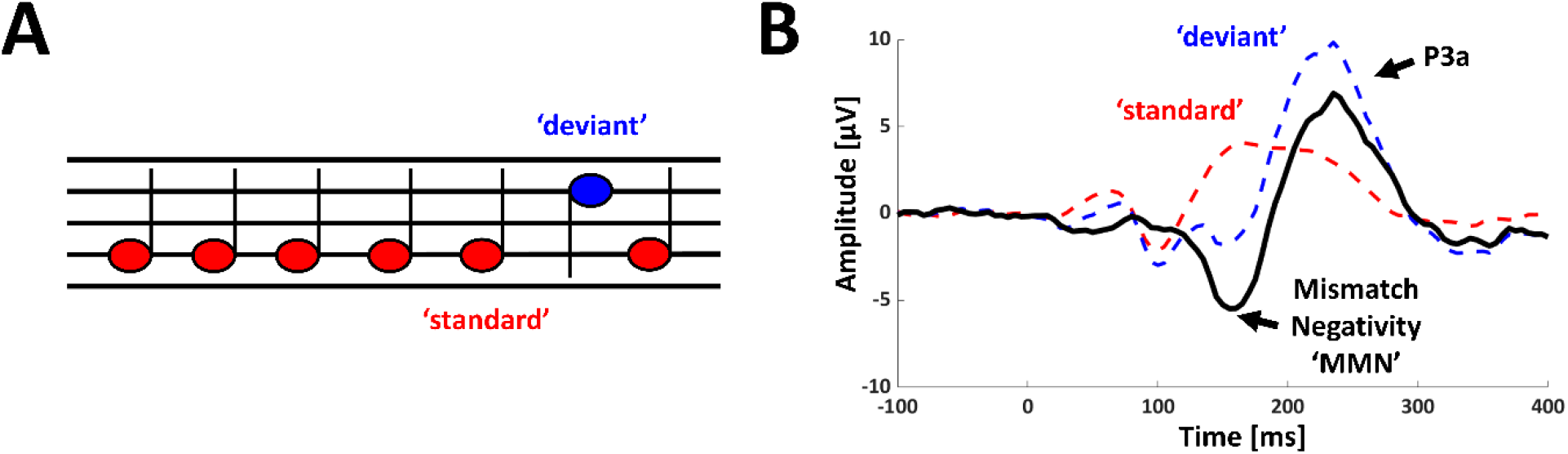
Mismatch negativity. **A** Example stimulus sequence to elicit auditory mismatch negativity. Violation of statistical regularity (established through repetition of low tones) elicits mismatch negativity. **B** Mismatch negativity is typically identified in the ERP difference wave obtained by subtracting response to the frequent and therefore more predictable tone (*standard*) from the response to the infrequent unpredictable tone (*deviant*).

MMN amplitude reductions have been replicated in numerous studies of patients with schizophrenia.^4^ A number of pharmacological challenge studies investigating the neuroreceptor basis of the MMN showed *N*-methyl-D-aspartate receptor (NMDAR) antagonists (e.g., phencyclidine or ketamine) to reduce MMN amplitude in rodents,^5^ monkeys,^6^ and humans^7–9^ (see ^10^ for an overview). Together, these results implicate NMDAR dysfunction as a pathophysiological mechanism underlying MMN amplitude reduction and a wide range of clinical symptoms and cognitive impairments in schizophrenia.^11–17^

Interest in MMN amplitude reductions has increased in recent years as a possible early warning sign for impending psychosis. MMN reductions are already present in individuals at clinical high-risk for psychosis (CHR-P), likely reflecting vulnerability for a progression to full psychosis as MMN amplitude reductions were found to be more pronounced in CHR-P individuals who later converted to a psychotic disorder.^18–22^ Despite its clinical potential, the mechanisms that account for these MMN alterations in the CHR-P population remain poorly understood.

One of the biggest challenges in early detection and intervention research lies in development of novel medications to delay or even prevent the transition to psychosis in CHR-P individuals.^23^ This challenge has been attributed to a lack of mechanistic models of pathophysiological processes, especially in the CHR-P population.^23^ In a previously published study, we found MMN amplitudes in ESZ patients and CHR-P individuals to be reduced,^21^ and further showed MMN deficits to be greater in those CHR-P individuals who subsequently converted to psychosis, relative to non-converters followed-up for at least 12 months. Here, we apply a computational approach^24^ to the EEG data from this prior study to develop a mechanistic model of altered information processing as a basis for MMN amplitude reductions in CHR-P individuals and ESZ patients.

## Methods and Materials

### Participants

Participants included 19 early-illness schizophrenia patients (ESZ; ≤5 years since initial hospitalization or initiation of antipsychotic medication), 38 CHR-P, and 44 healthy controls (HC).^21^ Clinical outcomes for the 38 CHR-P participants were tracked over 24 months, resulting in 15 CHR-P who converted to full psychosis and 16 CHR-P who did not convert but were followed for at least 12 months. Seven CHR-P dropped out before the 12-month follow-up and were, therefore, excluded from analyses comparing CHR-P converters and non-converters, since their clinical outcomes were less certain. ESZ participants were referred from the Yale Specialized Treatment Early in Psychosis (STEP) Clinic or from community clinicians. CHR-P were recruited from the Yale Psychosis Prodrome Research (PRIME) Clinic and HC through advertisements and word-of-mouth. The study was approved by the Institutional Review Board of Yale University and all adult participants provided written informed consent. For minors, parents provided written informed consent and minors provided written assent.

### Inclusion and exclusion criteria

ESZ patients met DSM-IV criteria for schizophrenia based on a Structured Clinical Interview for DSM-IV^25^ administered by a trained research assistant. CHR-P participants met the Criteria of Psychosis-Risk Syndromes based on the Structured Interview for Psychosis-Risk Syndromes (SIPS).^26,27^ Potential participants for all groups were excluded from the study if they fulfilled the following criteria: substance dependence or abuse within the past year, a history of significant medical or neurological illness or a head injury resulting in loss of consciousness, and abnormal audiometric testing. Additionally, HC who met criteria for any past or current DSM-IV Axis I disorder or had a first-degree relative with a psychotic disorder were excluded.

### Task

Participants performed an unrelated primary task (silently reading a book) while presented with three different auditory oddball paradigms presented in fixed order. Each paradigm comprised two runs of 875 tones each (1750 tones in total), including 90% standard tones (50ms, 633 Hz) and either 10% duration (100ms), 10% frequency (1000 Hz), or 10% duration + frequency double deviants (100ms *and* 1000 Hz). All tones were presented at 78 dB in fixed pseudorandomised order with 5ms rise/fall times and 510ms stimulus onset asynchrony through Etymotic ER3-A insert earphones (Etymotic Research, Inc., Elk Grove Village, Illinois).

### EEG data processing

EEG was recorded using a 20-channel electrode cap with a standard 10-20 montage (Physiometrix, Inc., North Billerica, Massachusetts) and additional mastoid and nose electrodes with linked-ear reference and an FPz ground. Signals were digitised at 1000 Hz with a Neuroscan Synamps amplifier (Neuroscan, Herndon, Virginia). Electro-oculograms were recorded from electrodes located above and below the left eye and at the outer canthi of both eyes.

EEG preprocessing consisted of high-pass filtering with a Butterworth filter (0.5 Hz), downsampling (256 Hz), low-pass filtering using a Butterworth filter (30 Hz), followed by epoching into 500ms segments around tone onsets (−100 to 400ms), baseline correction (−100 to 0ms), and eyeblink correction using principal component analysis with one component. Eyeblink components of all participants were manually inspected and eyeblink detection thresholds adjusted, if necessary, followed by rejection of remaining artefactual trials (using a ±100 μV amplitude threshold). Preprocessing and statistical analyses were implemented in MATLAB (version: 2020b; https://mathworks.com) using the SPM12 toolbox (https://www.fil.ion.ucl.ac.uk/spm/software/spm12/).

### Computational Modelling

We modelled implicit sensory learning about the tone sequences using a 3-level binary Hierarchical Gaussian Filter (HGF).^28,29^ This model assumes that participants make inferences about a number of hidden environmental states (Figure 2, left). In the context of the oddball paradigm, based on each trial input (*standard* or *deviant* tones), a participant needs to infer three hidden states, structured as follows: The lowest level state corresponds to the *tone probability*. On each trial *k* a tone can either be deviant 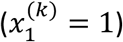 or a standard tone 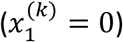. This state can be described by a Bernoulli distribution that is linked to the state at the second level *x*_2_^(k)^ through the unit sigmoid transformation:

**Figure 2.**
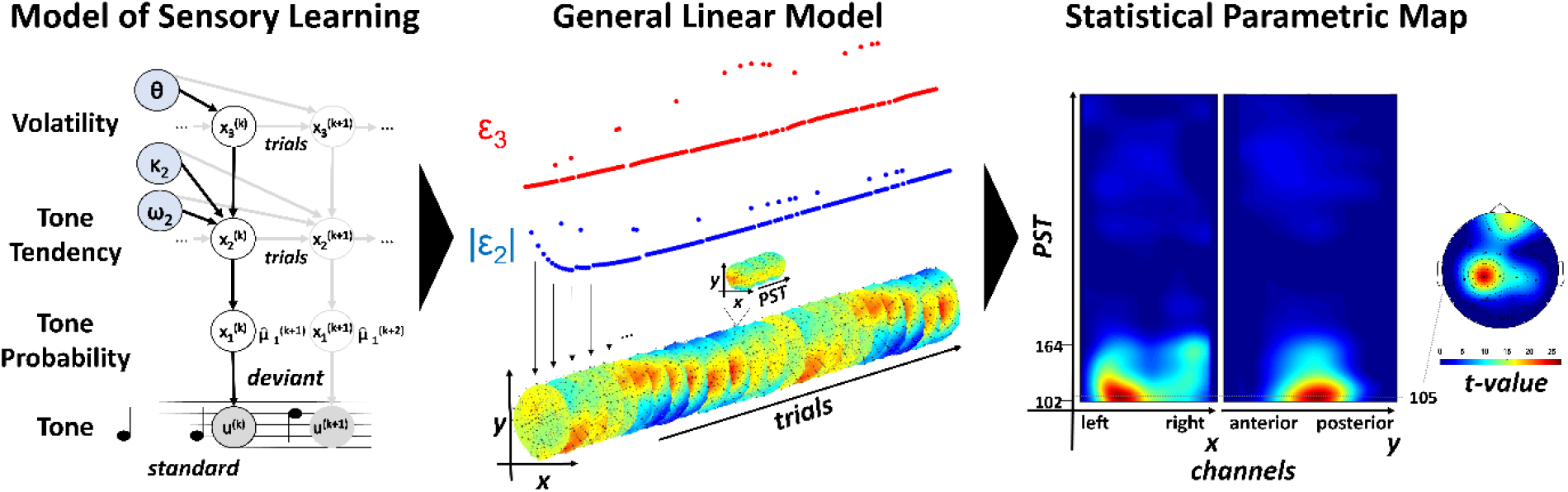
Computational analysis pipeline. Trial-by trial trajectories of low- and high-level precision-weighted prediction errors were computed using the Hierarchical Gaussian Filter^28,29^ (**left**). In a first level analysis, precision-weighted prediction errors were used as parametric regressors to explain EEG amplitude variations at each point in sensor space and peristimulus time (**PST**) following the tone presentation across trials within each participant (**middle**). First level statistics were carried to the second level to obtain statistical parametric maps over 2D sensor space and peristimulus time (**right**). **EEG**: Electroencephalography.

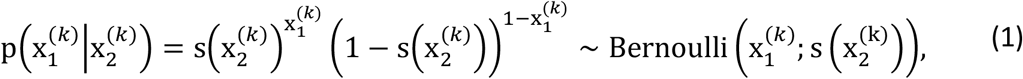

with

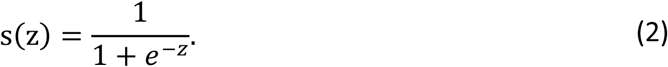

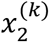 represents the unbounded tendency towards standard or deviant tones (−∞, +∞) or the *tone tendency* and is specified by a normal distribution:

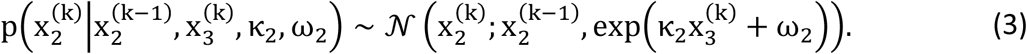

The state at the third level 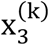 expresses the (log) volatility of the environment over time and is also modelled using a normal distribution:

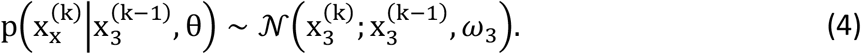

Participants’ beliefs about these hidden states at level *i* of the hierarchy and on trial *k* are denoted with 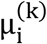 and updated after each new tone according to the following update equation:

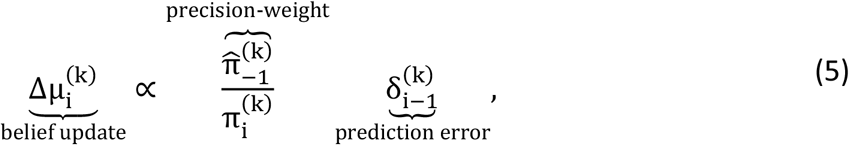

where 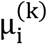 is the expectation or belief at trial *k* and level *i* of the hierarchy, 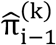 is the precision (inverse of the variance) from the level below (the hat symbol denotes that this precision has not been updated yet and is associated with the prediction before hearing a new tone), 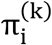 is the updated precision at the current level, and 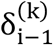 is a prediction error (PE) expressing the discrepancy between the expected and the experienced outcome.

In line with a previous study examining the effects of ketamine on sensory learning in a roving paradigm,^24^ we focused our analysis on low-level precision-weighted PEs about the tone tendency (ϵ_2_) and high-level precision-weighted PEs about the volatility of the environment (ϵ_3_), where the precision-weighted PE 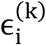 on each trial *k* and at level *i* of the hierarchy is defined as (cf. Eq. 5):

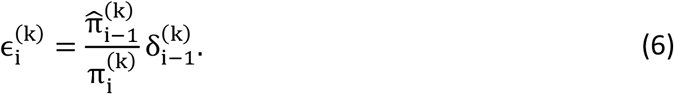

We implemented this model using the ‘tapas_ehgf_binary’ function from the HGF toolbox (version 6.0), which is made available as open-source code as part of the TAPAS^30^ software collection (version: 5.1.0; https://github.com/translationalneuromodeling/tapas/releases/tag/v5.1.0) in MATLAB (version: 2020b; https://mathworks.com). We used this recently developed enhanced version of the HGF to improve sensitivity to learning about environmental volatility. The main distinction with respect to earlier versions of the HGF is that the posterior means 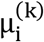 are updated before the precisions 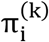 at level *i* of the hierarchy. For more details on the update equations, see ^28,29^ for the original HGF and the ‘tapas_ehgf_binary’ function for the eHGF.

We optimised the parameters of the perceptual model assuming that each participant acted as an ideal Bayesian observer minimising the surprise for a given input sequence using the ‘tapas_bayes_optimal_binary’ function. Ideally model parameters should be estimated based on both sensory input and participants’ behavioural responses to estimate how participants’ learning deviates from an ideal Bayesian observer. However, this was not possible, because the MMN paradigm is a passive task that does not require participants to make responses.

The prior settings (mean, variance) for this optimisation were (−3, 4) for the *evolution rate* ω_2_ and (2, 4) for meta-volatility ω_3_. The *coupling strength* κ_2_ was fixed to log(1). Posterior parameter estimates are summarised in Table 1.

**Table 1.** Summary of posterior parameter estimates.

|  | **HC**<br>*n*=44 | **CHR-P**<br>*n*=38 | **ESZ**<br>*n*=19 | **CHR-C**<br>*n*=15 | **CHR-NC**<br>*n*=16 |
| --- | --- | --- | --- | --- | --- |
| **evolution rate $\omega_2$**<br>mean [SD] | -0.20<br>[0.78] | -0.19<br>[0.77] | -0.36<br>[0.85] | -0.16<br>[0.82] | -0.22<br>[0.78] |
| **meta-volatility $\omega_3$**<br>mean [SD] | 4.80<br>[0.30] | 4.86<br>[0.29] | 4.80<br>[0.31] | 4.76<br>[0.29] | 4.83<br>[0.31] |

**HC:** Healthy controls. **CHR-P:** Individuals at clinical high risk for psychosis. **ESZ:** Early illness schizophrenia patients ( $\leq 5$ years since initial hospitalization or initiation of antipsychotic medication). **CHR-C:** Converters. **CHR-NC:** Non-converters

### Statistical analyses

Demographic and clinical variables were analysed in R (version: 4.04; https://www.r-project.org/) using R-Studio (version: 1.4.1106; https://www.rstudio.com/). We report uncorrected *p*-values for either ANOVAs or χ^2^-tests where appropriate. Post hoc tests were Bonferroni-corrected (α = 0.05).

#### First level analysis

We extracted the trajectories of low-level precision-weighted PEs about the tone tendency ϵ_2_ and high-level precision-weighted PEs about the volatility of the environment ϵ_3_. Trial-by trial magnitude estimates of the absolute value of low-level precision-weighted PEs |ϵ_2_| or high-level precision-weighted PEs ϵ_3_ were included as parametric regressors to explain trial-by-trial variation in EEG amplitude (Figure 2) as done previously.^24^ The absolute value of ϵ_2_ was chosen because it expresses Bayesian surprise independent of the physical characteristics of a tone such as a specific frequency. The general linear model at the first level consisted of an intercept term and either (z-standardized) low or high-level precision-weighted PE trajectories as predictors and EEG amplitude across sensors and peri-stimulus time as the response variable. For each precision-weighted PE, we tested the null hypothesis that the parameter estimate was zero at each sensor and time point using an F-test. Statistical analyses were restricted to 100 to 400ms post-stimulus time.

#### Second level analysis

First-level statistics were converted into images and smoothed using a Gaussian kernel (FWHM: 16 mm x 16 mm) to ensure that the assumptions of Gaussian random field theory were met.^31,32^ Smoothed images were carried to the second level to compare groups using different factorial designs for each precision-weighted PE to obtain statistical parametric maps over 2D sensor space and peri-stimulus time (Figure 2). Each factorial design included group as between- and MMN oddball paradigm as within-subject factor, as well as age as a covariate. To ensure that the equal slope assumption for age was met, we masked out voxels that showed a significant group-by-age interaction. Multiple testing correction was implemented using Gaussian random field theory^31,32^ and we report *p*-values corrected for peak-(*ppFWE*) or cluster-level (*pcFWE*) family-wise error rates using a cluster defining threshold of *p*<0.001^33^ unless stated otherwise.

## Results

### Demographic and clinical characteristics

Demographic and clinical characteristics are displayed in Table 2 (see also ^21^ for more information).

**Table 2.**
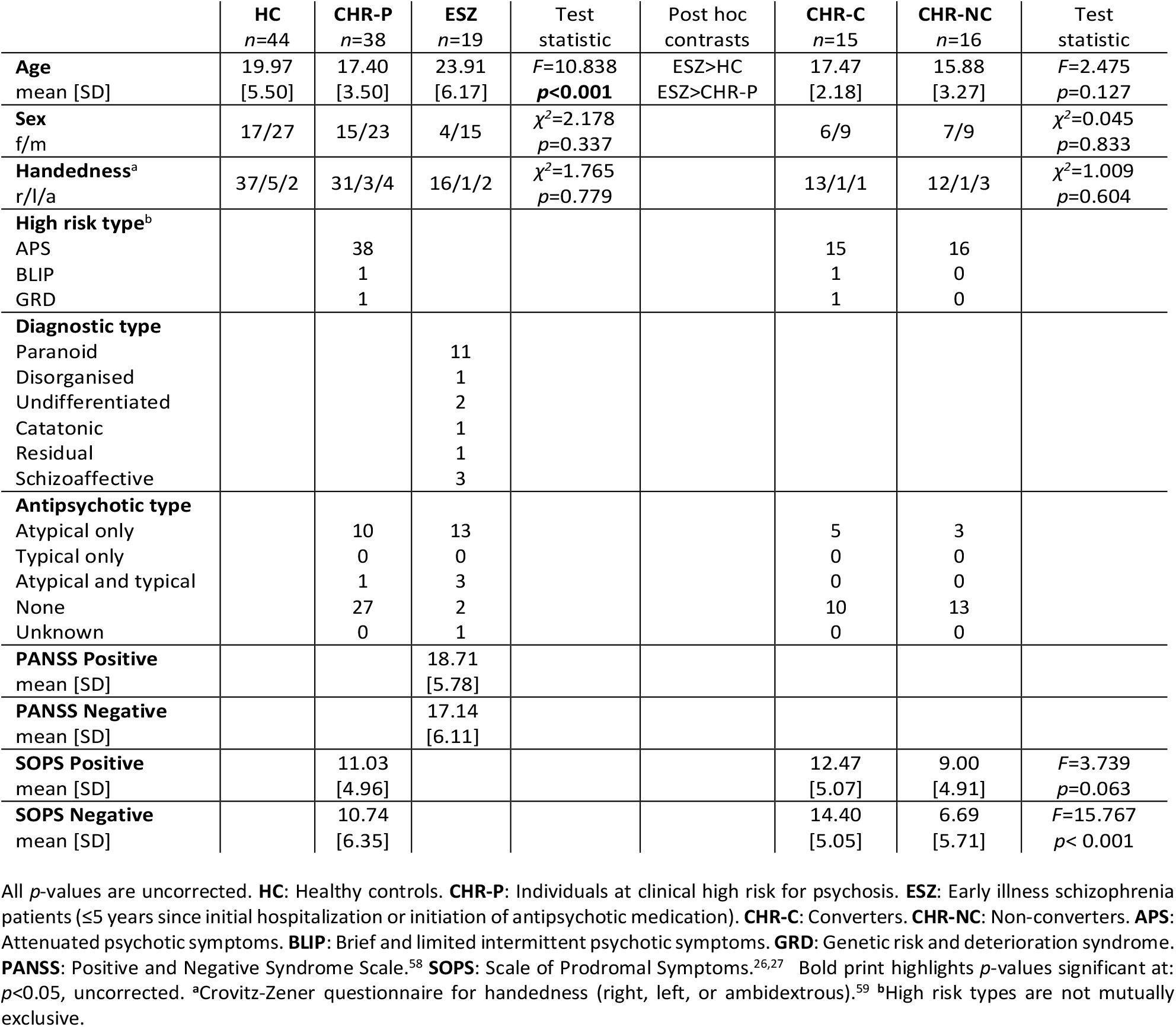
Demographic and clinical characteristics.

### Group differences in low-level precision-weighted prediction errors

We observed a significant group effect on the expression of low-level precision-weighted PEs about the tone tendency ϵ_2_ peaking at 105ms over left, central channels (*F*=20.795, *pcFWE*<0.001) and at 109ms over frontal channels (*F*=15.656, *ppFWE*<0.001). Closer inspection of the first effect revealed that the difference between small and large low-level precision-weighted PEs was reduced in central channels in ESZ compared to CHR-P (peak: 152ms, *t*=4.923, *pcFWE* =0.001; Figure 3) and ESZ vs HC (peak: 105ms, *t*=6.427, *pcFWE*<0.001; Figure 3). The second effect again suggested a reduced difference between small and large low-level precision-weighted PEs. However, this effect was expressed over frontal channels in ESZ vs HC (peak: 109ms, *t*=5.594, *ppFWE*<0.001; Figure 3). The timing of these effects coincided with the timing of the MMN^21^ suggesting that MMN reductions may reflect disturbances in precision-weighted PE updating processes as hypothesised previously.^34,35^

**Figure 3.**
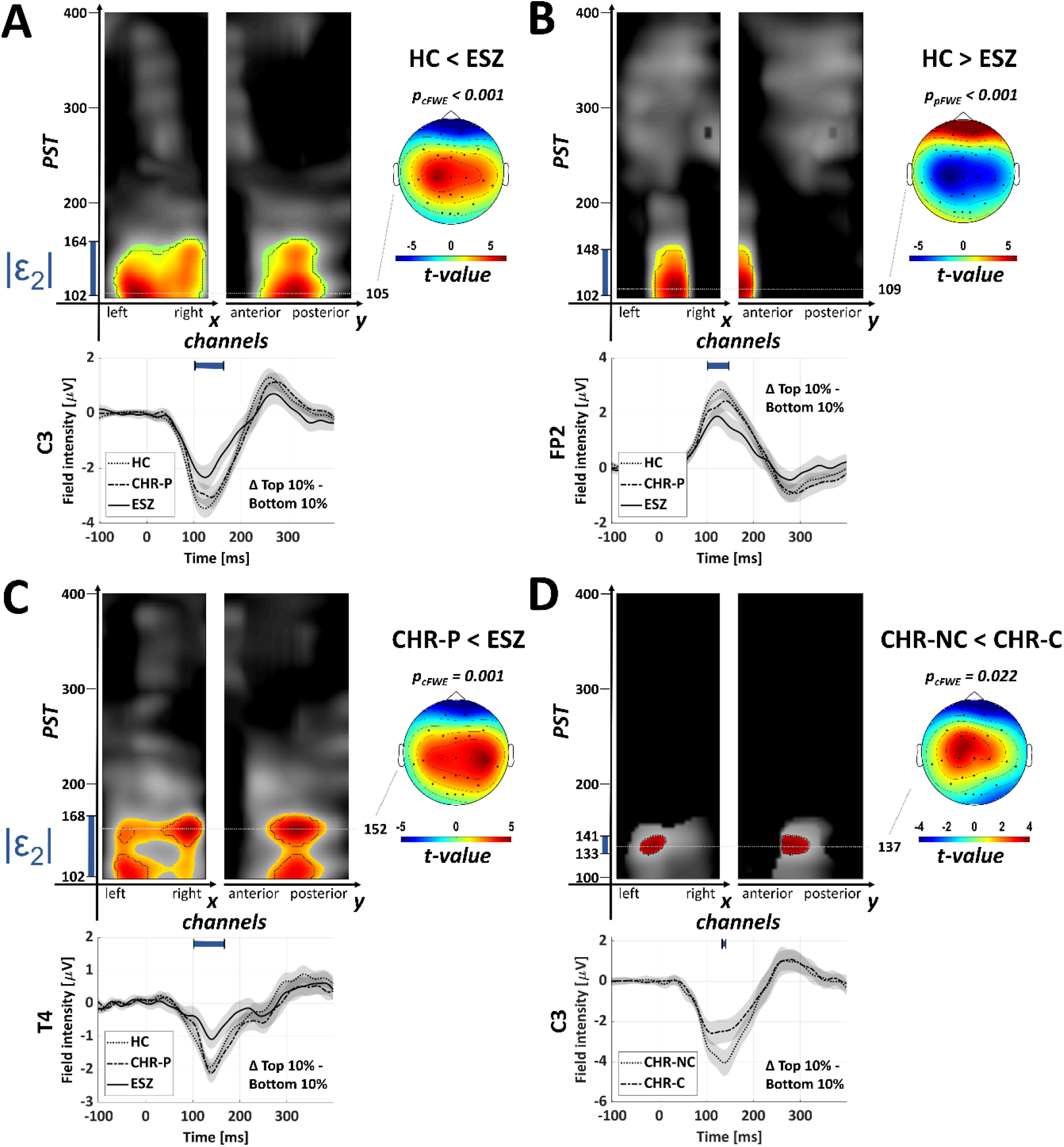
Low-level precision-weighted prediction errors *ε*_2_. **A-D**: Displayed are maximum intensity projections highlighting significant voxels of *t*-contrasts testing for pairwise group differences in the expression of low-level precision-weighted prediction errors *ε*_2_ about the tone tendency. Times displayed on y-axis indicate earliest and latest significant voxel. *p*-values were corrected for peak-(*p*_*pFWE*_; black dashed-line) or cluster-level (*p*_*cFWE*_) family-wise error rates (**FWE**) using a cluster defining threshold of *p*<0.001 (highlighted by coloured area). Note, that *p*-values in **D** are small-volume corrected for the group effect on *ε*_2_ between HC and ESZ (i.e., significant voxels in plots **A** OR **B**; black colour illustrates masked out voxels in **D**). For illustration, difference waveforms (10% highest −10% lowest *ε*_2_ trials) are shown across groups for a channel close to the peak effect. **HC**: Healthy controls. **CHR-P**: Individuals at clinical high risk for psychosis. **CHR-C**: Converters. **CHR-NC**: Non-converters. **ESZ**: Early-illness schizophrenia patients (≤5 years since initial hospitalization or initiation of antipsychotic medication). **PST**: Peristimulus time following tone presentation. Note that the statistical analysis window was restricted to 100-400ms following each tone (standard and deviants).

### Group differences in high-level precision-weighted prediction errors

The expression of high-level precision-weighted PEs about the volatility of the environment ϵ_3_ also showed a significant effect of group peaking at 125ms over right, central channels (*F*=17.277, *pcFWE*=0.005). Pairwise comparisons revealed stronger correlations of high-level precision-weighted PEs with EEG amplitudes in HC compared to ESZ over frontal channels (peak: 125ms, *t*=3.931, *ppFWE*=0.027) and during a later time window over posterior central channels (peak: 344ms, *t*=3.821, *pcFWE*=0.018; Figure 4), which was also significant when comparing CHR-P to ESZ (peak: 340ms, *t*=3.621, *pcFWE*=0.046; Figure 4). Furthermore, we found that the difference between small and large precision-weighted PEs was reduced during an early time window in ESZ vs CHR-P (peak: 129ms, *t*=5.014, *pcFWE*=0.008; Figure 4) and in ESZ vs HC (peak: 125ms, *t*=5.728, *pcFWE*=0.002; Figure 4). While the early cluster again coincided with the time window of the MMN^21^; the latter cluster rather fell into the P3a time window, raising the question whether the P3a may also reflect PE-related processing (cf. Figure 1).

**Figure 4.**
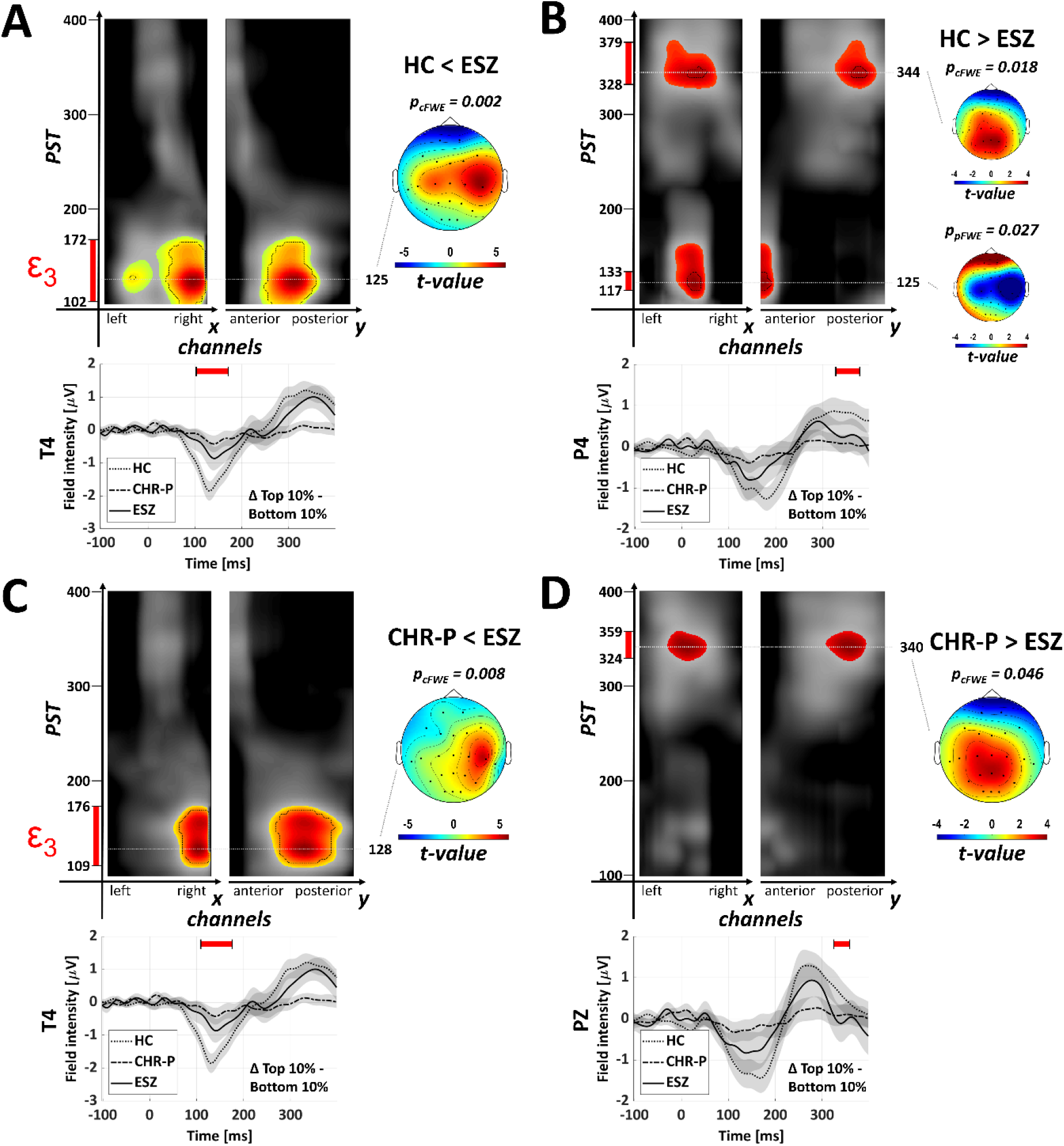
High-level precision-weighted prediction errors. **A-D**: Displayed are maximum intensity projections highlighting significant voxels of *t*-contrasts testing for pairwise group differences in the expression of high-level prediction errors ϵ_3_ about environmental volatility. Times displayed on y-axis indicate earliest and latest significant voxel. *p*-values were corrected for peak-(*p*_*pFWE*_; black dashed-line) or cluster-level (*p*_*cFWE*_) family-wise error rates (**FWE**) using a cluster defining threshold of *p*<0.001 (highlighted by coloured area). For illustration, difference waveforms (10% highest −10% lowest *ε*_3_ trials) are shown across groups for a channel close to the peak effect. **HC**: Healthy controls. **CHR-P**: Individuals at clinical high risk for psychosis. **ESZ**: Early-illness schizophrenia patients (≤5 years since initial hospitalization or initiation of antipsychotic medication). **PST**: Peristimulus time following tone presentation. Note that the statistical analysis window was restricted to 100-400ms following each tone (standard and deviants).

### Group differences between converters and non-converters

Lastly, when comparing CHR-P converters to non-converters, we found a significant group effect on the expression of low-level precision-weighted PEs ϵ_2_ peaking at 137ms over left, central channels (*F*=12.722, *pcFWE*=0.040; small-volume corrected for the group effect on ϵ_2_ between HC and ESZ). In CHR-P individuals who later transitioned to psychosis, the difference between small and large low-level precision-weighted PEs was reduced (peak: 137ms, *t*=3.567, *pcFWE*=0.022; small-volume corrected for the group effect on ϵ_2_ between HC and ESZ; Figure 3D).

## Discussion

The objective of this study was to develop and test a mechanistic model of altered information processing as a basis for MMN amplitude reductions in CHR-P individuals and ESZ patients. We obtained three major findings: First, we observed altered expression of low-level precision-weighted PEs about the tone tendency between HC and ESZ and in CHR-P compared to ESZ. Second, we also identified changes in the expression of high-level precision-weighted PEs about the volatility of the environment in ESZ compared to both HC and CHR-P during an early time window (at about 100-175ms peristimulus time), as well as during a later time window (at about 320-380ms). Third, the expression of low-level precision-weighted PEs was significantly altered in those CHR-P that later converted to a psychotic disorder compared to non-converters suggesting that this computational model appears to capture relevant pathophysiological mechanisms and may constitute a useful tool to predict transition to psychosis in CHR-P individuals.

### Theoretical implications for the predictive coding account of psychosis

Our results are in line with the predictive coding account of psychosis that postulates that disturbances in hierarchical PE processing may contribute to psychotic symptoms.^14,36^ Our finding of alterations in the expression of precision-weighted PEs in central channels in patients with schizophrenia may suggest that patients experience aberrantly salient PEs^37^ in response to familiar stimuli (*standard* tones). Furthermore, changed expression of hierarchical PEs in frontal channels could signal a decrease in the precision of priors in frontal regions, as proposed in the predictive coding account of psychosis.^14^

### Possible cortical generators of aberrant prediction errors

The network of cortical regions thought to be involved in generating the MMN response includes bilateral primary auditory cortices (A1), superior temporal gyri (STG) and inferior frontal gyri (IFG).^34,38–40^ It is possible that the different spatiotemporal clusters that were identified in our study may be caused by different cortical generators, for example the correlation between precision-weighted PEs and EEG amplitudes in ESZ expressed in central channels may originate in A1 or STG, while the second cluster, which was identified over frontal regions, possibly suggests involvement of IFG. However, due to volume-conduction effects, we will have to formally test this hypothesis using source modelling in the future. Adams and colleagues^41^ recently investigated the neural mechanisms of schizophrenia using dynamic causal modelling. They found remarkably consistent findings across a wide range of paradigms implicating reductions in a model parameter that serves to amplify the magnitude of activity or *synaptic gain* of pyramidal cells. Notably, this study also included an MMN paradigm, in which the authors identified reduction of pyramidal gain in IFG specifically. This finding suggests that the expression of hierarchical PEs over frontal channels may be altered due to a reduction of the precision-weight rather than changes in the PE component of the precision-weighted PEs.

### Are aberrant precision-weighted prediction errors related to alterations in neurotransmission?

The dysconnectivity hypothesis^13,16,42–44^ postulates that NMDAR-mediated modulation of synaptic gain is altered in schizophrenia. In line with this account, Weber et al.^24^ found that ketamine administration led to a reduced expression of high-level precision-weighted Pes about the volatility of the environment in central channels, similar to our results. However, their results suggest that low-level precision-weighted PEs are unaffected by ketamine.

Several neurotransmitters interact with NMDAR to dynamically control synaptic gain and neuroplasticity. Altered expression of precision-weighted PEs in ESZ, as identified in our study over early auditory regions, could reflect changes in cholinergic neurotransmission. Two recent studies implicate acetylcholine in regulating synaptic gain or — according to the predictive coding account and the dysconnectivity hypothesis — regulating sensory precision in early auditory regions.^35,45^ The first study employed a Kalman filter (i.e., a 2-level HGF) to model changes in participants that were administered galantamine, which enhances cholinergic neurotransmission.^35^ The authors argued that galantamine may increase the precision of sensory PEs. The second study by Schöbi and colleagues^45^ modelled changes in between- and within-region connectivity including synaptic gain during a pharmacological manipulation using muscarinic receptor antagonist scopolamine or muscarinic receptor agonist pilocarpine in rats.^45^ The authors found dose-dependent changes in synaptic gain, but also changes in inter-regional connectivity between A1 and secondary auditory cortex. Moreover, changes in muscarinic receptor density among schizophrenia patients have been frequently reported ^46–49^ and Scarr et al.^48^ proposed that there may be a subgroup of schizophrenia patients specifically characterized by decreased cortical muscarine receptor expression. These results support a potential role of cholinergic neurotransmission in precision-weighted PE signalling. Beyond cholinergic processes, glutamatergic neurotransmission at AMPA receptors may be involved, but its precise role still needs to be clarified.

### Clinical Implications

Interestingly, our results suggest that the expression of low-level precision-weighted PEs is blunted in CHR-P that later converted to a psychotic disorder compared to CHR-P that did not convert. This finding highlights potential applications of this computational approach to prediction of psychosis in CHR-P individuals. Furthermore, if the neurotransmitter systems that are involved in computing precision-weighted PEs during the MMN paradigm can be identified, this approach may be useful for identifying critical time windows for preventative interventions or for predicting treatment response to pharmacological interventions that target either glutamatergic neurotransmission like d-serine, which has shown promising results in a recent clinical trial,^50^ or cholinergic neurotransmission, for example involving muscarinic (M1, M4) agonist xanomeline.^51^

### Limitations

A few limitations of this study merit attention. First, the oddball paradigms in this study were not well-suited to separate low- and high-level PEs because environmental volatility was not manipulated explicitly throughout the task. Future studies should include explicit manipulations of volatility (i.e., changes in deviant probabilities over the course of the experiment, rather than a 90% stable probability) to better distinguish between different levels of hierarchical inference. Secondly, we assumed that participants acted as ideal Bayesian observers without taking subject-specific deviation from an ideal Bayesian observer into account by estimating subject-specific parameters based on both sensory input *and* behavioural responses. This is an inherent limitation of the *passive* MMN oddball paradigm. In light of this limitation, our results are more challenging to interpret. Group differences could arise because different groups are better explained by different models or model parameter values. While MMN amplitude reductions have been frequently replicated in schizophrenia,^4^ future studies should also investigate the representation of precision-weighted PEs using *active* oddball paradigms that require participants to detect and respond to infrequent target stimuli, such as the paradigm used in a recent study,^52^ which found that target P3b amplitudes were predictive of conversion to psychosis.

### Future directions

Future studies should determine the cortical generators of changes in the expression of hierarchical precision-weighted PEs in the clinical high risk for psychosis state and early schizophrenia. Moreover, the biological implementation of these computations needs to be clarified further, for example through the use of models that include greater physiological detail to bridge the algorithmic description that our modelling approach offers and its physiological implementation in the brain.^53^ Dynamic causal models for electrophysiological data have been highlighted as computational assays that may allow to infer receptor densities of neuronal populations.^13,54^ These models have been validated in studies investigating NMDAR antibody encephalitis,^55^ dopaminergic action on NMDARs,^56^ and manipulations of cholinergic neurotransmission,^45^ and thus, constitute a promising way forward. Additionally, there is a need for more pharmacological studies in both animals and humans to map the relationship between hierarchical precision-weighted PEs and different neurotransmitter systems that are targeted by antipsychotic medication.

### Conclusions

In this study, we examined the computational mechanisms underlying pre-attentive auditory deviance processing in the clinical high risk for psychosis state and early schizophrenia and found evidence for aberrant expression of precision-weighted PEs at different levels of hierarchical inference. Our results suggest that the expression of low-level precision-weighted PEs is significantly altered in individuals at clinical high risk for psychosis that will later transition to psychosis, highlighting that this computational modelling approach captures relevant pathophysiological mechanisms and may prove useful for predicting transition to psychosis in CHR-P individuals.

## Data Availability

Data sharing is not applicable to this article as no new data were created or analyzed in this study.

## Acknowledgements

This work was supported by the National Institutes of Health (R01 MH076989 to DHM), the Swiss National Science Foundation (Doc.Mobility, 200054 to DJH; Ambizione, PZ00P3_167952 to AOD), the Brain and Behavior Research Foundation (to DHM), the Krembil Foundation (to AOD), and the U.S. Department of Veterans Affairs (Veterans Affairs Senior Research Career Award, 1IK6CX002519 to JMF).

A previous version of this article was published as part of DJH’s PhD thesis^57^ and made available as a preprint on https://www.medrxiv.org/.

